# Personalized labeling: A strategy for supporting self-medicating patients’ decision-making during selection and use

**DOI:** 10.1101/2025.08.01.25332697

**Authors:** Lanqing Liu, Mark Becker, Sukhdeep Singh, Dangkamol Wongthanaroj, Laura Bix

## Abstract

Interactions between self-medicating consumers and labeling of Over-the-Counter medications (OTC) influence the quality of information processing and, hence, appropriateness of decisions. Our previous work yielded evidence that during the early stages of information processing important regulatory information was frequently unnoticed and that decision-making related to OTCs could be improved.

We postulated the concept of “personalized labeling” would address recognized shortcomings affiliated with current OTC labels. sThis strategy uses an augmented reality interface to present users with a recommendation related to an OTC’s appropriateness for the individual’s use while the product is being viewed through their smart phone. In doing so, it advises decisions specific to the user’s health history and medication usage, assisting with appropriate decision making.

To develop data in support of a proof of concept for this strategy, we utilized an *absolute judgement test.* Seventy-two participants divided into two cohorts (educated to the personalized labeling strategy vs control) made binary decisions related to a drug’s appropriateness for use by a theoretical patient considering single-ingredient products. Response variables measured the approach’s effectiveness (response accuracy) and efficiency (time to accurate decision).

When educated to the personalized labeling concept, participants made decisions significantly more accurately (personalized: ME=0.977, SE=0.007; standard: ME=0.933, SE=0.017; p=0.002) and faster (personalized: ME=9.584s, SE=0.854; standard: ME=19.052s, SE=2.322; p<0.001) for trials with personalized Front-of-Pack (FOP) labels compared to trials composed of the current commercial standard.

This suggests that personalized labeling has the potential to improve consumer decision making related to OTC selection and use. Future studies are needed utilizing a broader range of populations, package types, and use contexts.

## Introduction

Beginning in the 1990s, a Drug Facts Label (DFL), a comprehensive, standardized label containing critical information specific to medications sold over the counter, became required on most over the counter (OTC) medications sold in the US. DFLs are intended to provide a systematized, consumer-focused way to display information critical for the safe and effective use of OTC products at the points of purchase and use. Effective labeling is particularly important for OTC medications, as they are deemed to be safe and effective absent a healthcare provider’s supervision *when used as directed*.

That said, DFLs have been criticized through the years. Critiques of the existing approach include: the use of small print (1,2) and crowded formatting with extraneous information present (1); complex wording (2,3) that results in poor comprehension; a tendency of consumers to preferentially attend marketing information (4–7); and designs that are not optimized for those with poor health literacy (8–10) or who speak English as a second language (11,12). In short, the DFL needs to be improved.

Recognizing these critiques, Catlin and Brass (2018) conducted an analysis of the existing literature related to DFL design. The authors conclude that even DFL designs that perform “reasonably well” in comprehension studies (required by the US Food and Drug Administration (FDA)) do not communicate effectively in studies that are more complex or multifaceted, such as self-selection or actual use studies. This led the authors to conclude that comprehension studies should be viewed as “idealized estimates” of DFL performance. They indicate the “static and non-customizable" nature of current DFLs prevent them from “uniformly meeting the needs” of several populations, including: those with limited literacy, the visually impaired, people with language barriers, older consumers and those with pre-existing believes and attitudes that override DFL messaging (3). The authors also hypothesize that the multi-attribute nature of information processing complicates decision making and creates difficulty for consumers who struggle to integrate many factors to arrive at an appropriate, specific decision.

We theorize that the use of an app containing the health history and medication use of an individual which employs augmented reality to return a customized response related to an OTC’s appropriateness for the app owner’s use would transcend the shortcomings of traditional printing, addressing many of the concerns identified by Catlin and Brass’ (2018), and, therefore, better serve the vulnerable audiences they highlight. Under such a paradigm, a consumer would “interrogate” a potential medication utilizing visual recognition technology in their smart device (3). The device would recognize the product, cross reference any contraindications from the label with the health and medication history loaded in the app, and return a customized response (yes, OK; no, contraindicated) using augmented reality, which “floats” the response above the interrogated product. (See Fig 1)

**Fig 1.**
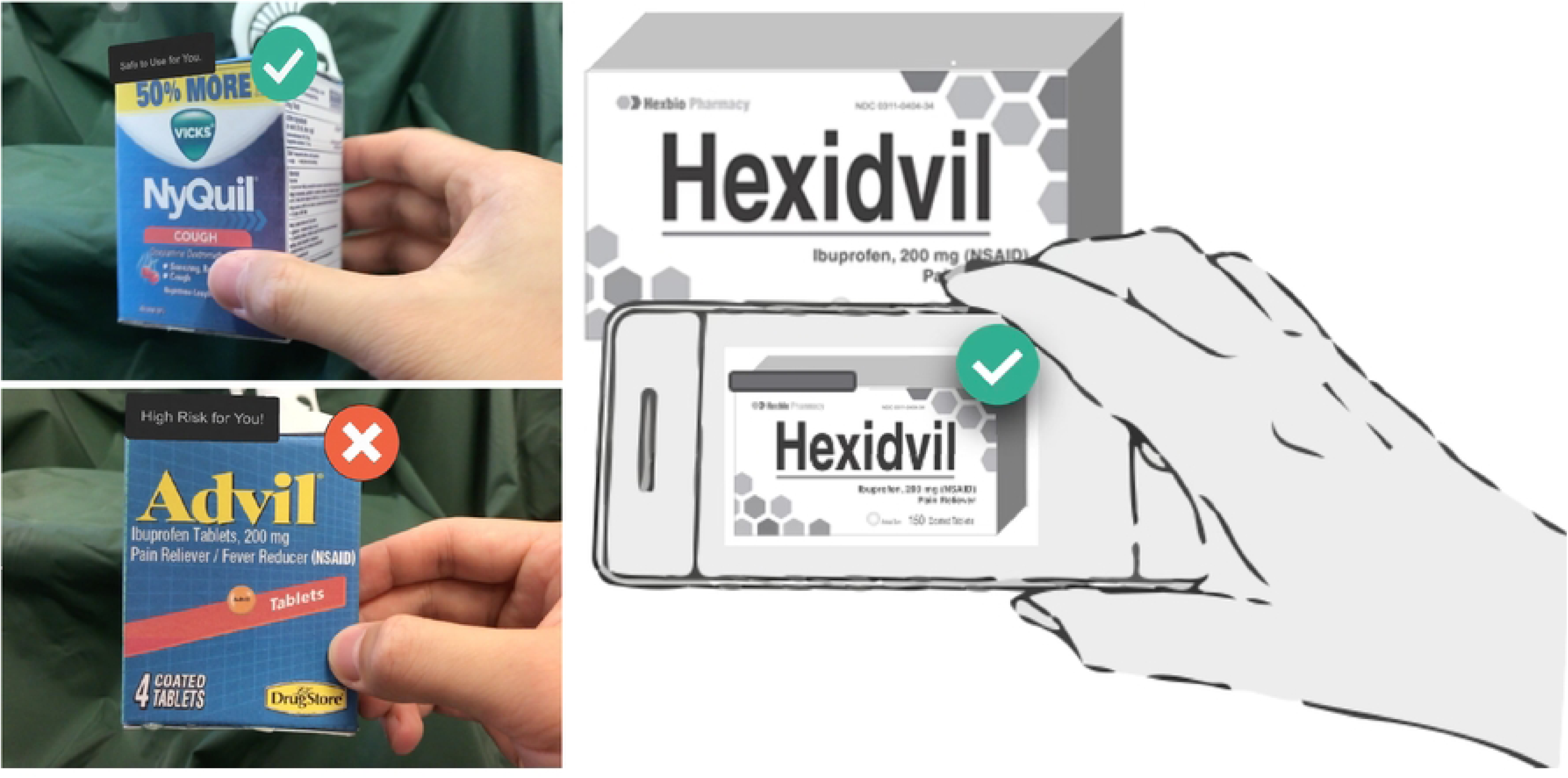
Personalized labeling strategy with the use of augmented reality.

To sum, we envision a label that is flexible, or “personalized”, which adapts on a case-by-case basis by cross referencing the drug under consideration to information individual to the person deliberating its use.

Herein, we evaluate the potential benefits personalized labeling could provide in terms of effectiveness (the accuracy of choice) and efficiency (the resources it takes to make that choice, measured as time to a correct decision). To obtain these dependent variables, we performed a computer-based, absolute-judgement task using scenarios where participants were asked whether a given OTC was appropriate to use by an individual in a prepared scenario (yes/no).

### Objectives

The overarching objective of our research is to enhance OTC labeling in ways that address problematic issues that are well-documented in the literature. Herein we develop data to support personalized labeling as a proof of concept. We frame this experiment using ISO/IEC 9241, “Ergonomics of human-system interaction-Part 210: Human-centered design for interactive systems.” The ISO standard suggests that usability is a confluence of three factors: (*1*) *effectiveness,* the “accuracy and completeness with which users achieve specified goals,” (*2*) *efficiency,* the “resources used in relation to the results achieved,” and (*3*) *satisfaction*, the “extent to which the user’s physical, cognitive and emotional responses that result from the use of a system, product or service meet the user’s needs and expectations.”

We assess two of the three usability metrics; namely, the effectiveness (assessed by the proportion of correct responses) and efficiency (assessed as the time it took respondents to record a correct response) of the new, theorized approach to labeling. Each dependent variable allows us to compare the DFL used in commercial practice to the novel strategy (personalized labeling).

## Material and methods

### 1. Participants and recruitment

Procedures were conducted in accordance with approvals provided by the Michigan State University Office of Regulatory Affairs Human Research Protection Program (STUDY00005057) as exempt.

Power estimates for this study were based on previous work (13) suggesting an effect size (d) of d=0.84. The previous work focused on surgical technicians and healthcare providers who were required to select appropriate medical devices for specific scenarios (e.g. latex allergic patient) as part of their job (surgical technologists); the power estimate was conducted with 30% of the measured effect size for this study which employed a more general population, or d=0.25. It was determined that 66 participants were estimated as allowing us to detect d=0.25 with given α=0.05 and power >0.8.

Seventy-two participants were recruited via the SONA recruiting system available from the MSU College of Communication Arts and Sciences, distribution of IRB approved flyers, and word of mouth. Participants were eligible for this study if they were: legally sighted, 18 years or older, had used OTC medications within 6 months of the experiment, and had transportation to campus, where the study took place. Participants were characterized using a brief demographic survey followed by tests of their: (1) Near-point visual acuity; (2) Adult Literacy in Medicine (measured with the revised REALM-R; and (3) Color differentiation test.

Half of the participants (n=36) were assigned to a “concept-educated” group and half (n= 36) the control group (see Table 1 for demographic information). Participants in the concept-educated group were informed that the test was meant to explore the usability of an application that utilized augmented reality, returning a customized response regarding appropriateness for those who operated an app on their smart device. They were also informed about the meaning of the “checkmark” symbol and the red “stop sign” symbol. Participants in the control group were not provided any information about the concept or the use of symbols.

**Table 1.** Frequency Table- Participants across Demographic Factors by Group Assignment.

| **Characteristic** | **Group Assignment** | **Value** | **Number** | **% of Total (72)** |
| --- | --- | --- | --- | --- |
| **Sample size** | **Concept Educated** | N1 | 36 | 50% |
|  | **Control** | N0 | 36 | 50% |
| **Sex** | **Concept Educated** | Male | 8 | 11.1% |
|  |  | Female | 28 | 38.9% |
|  | **Control** | Male | 11 | 15.3% |
|  |  | Female | 25 | 34.7% |
| **Color Differentiation** | **Concept Educated** | Normal | 36 | 50.0% |
|  |  | Color blind | 0 | 0.0% |
|  | **Control** | Normal | 35 | 48.6% |
|  |  | Color blind | 1 | 1.4% |
| **Education** | **Concept Educated** | Masters or higher | 14 | 19.4% |
|  |  | Bachelor or lower | 22 | 30.6% |
|  | **Control** | Masters or higher | 8 | 11.1% |
|  |  | Bachelor or lower | 28 | 38.9% |
| **Ethnicity** | **Concept Educated** | White | 22 | 30.6% |
|  |  | Others | 14 | 19.4% |
|  | **Control** | White | 24 | 33.3% |
|  |  | Others | 12 | 16.7% |
| **Health Literacy** | **Concept Educated** | Normal | 35 | 48.6% |
|  |  | Risk for poor literacy | 1 | 1.4% |
|  | **Control** | Normal | 35 | 48.6% |
|  |  | Risk for poor literacy | 1 | 1.4% |
| **Native Language** | **Concept Educated** | English | 27 | 37.5% |
|  |  | Others | 9 | 12.5% |
|  | **Control** | English | 30 | 41.7% |
|  |  | Others | 6 | 8.3% |
| **Visual Acuity** | **Concept Educated** | Normal ($\leq 20/40$) | 36 | 50.0% |
| | | Poor ($>20/40$) | 0 | 0.0% |
| | **Control** | Normal ($\leq 20/40$) | 36 | 50.0% |
| | | Poor ($>20/40$) | 0 | 0.0% |
| **Age** | **Concept Educated** | Mean (Min, Max) | 34.64 (19, 65) |  |
|  |  | Std. Deviation | 13.226 |  |
|  | **Control** | Mean (Min, Max) | 34.19 (20, 73) |  |
|  |  | Std. Deviation | 14.903 |  |

### 2. Stimuli and trials

A custom-built program (E Prime 3.0 Tools Psychology Software; Pennsylvania USA) was used to create an absolute judgment task which recorded both correct and incorrect responses and response time. Eight single-ingredient products were utilized as the basis for the test trials. Each of the eight products was depicted once with a scenario where the correct response was yes and a second time where the correct response was no. Response was crossed with two label treatment at two levels (standard formatting (with the traditional DFL) and the personalized label), a total of 4 trials per active ingredient (See Fig 2 for a single set affiliated with one product), for a total of 32 test trials (8 products × 2 labeling presentations (standard/personalized) × 2 answer types (yes-appropriate/no- not appropriate)). The following active ingredients were tested: acetaminophen (ACE— scientifically referred to as APAP) – Tylenol; ibuprofen (IBU) – Advil; naproxen (NAP) – Aleve; guaifenesin (GUA) - Mucinex, omeprazole (OME) – Prilosec; phenylephrine (PHN-scientifically referred to as PE) – Sudafed; cimetidine (CIM) – Tagamet; ranitidine (RAN) – Zantac.

**Fig 2.**
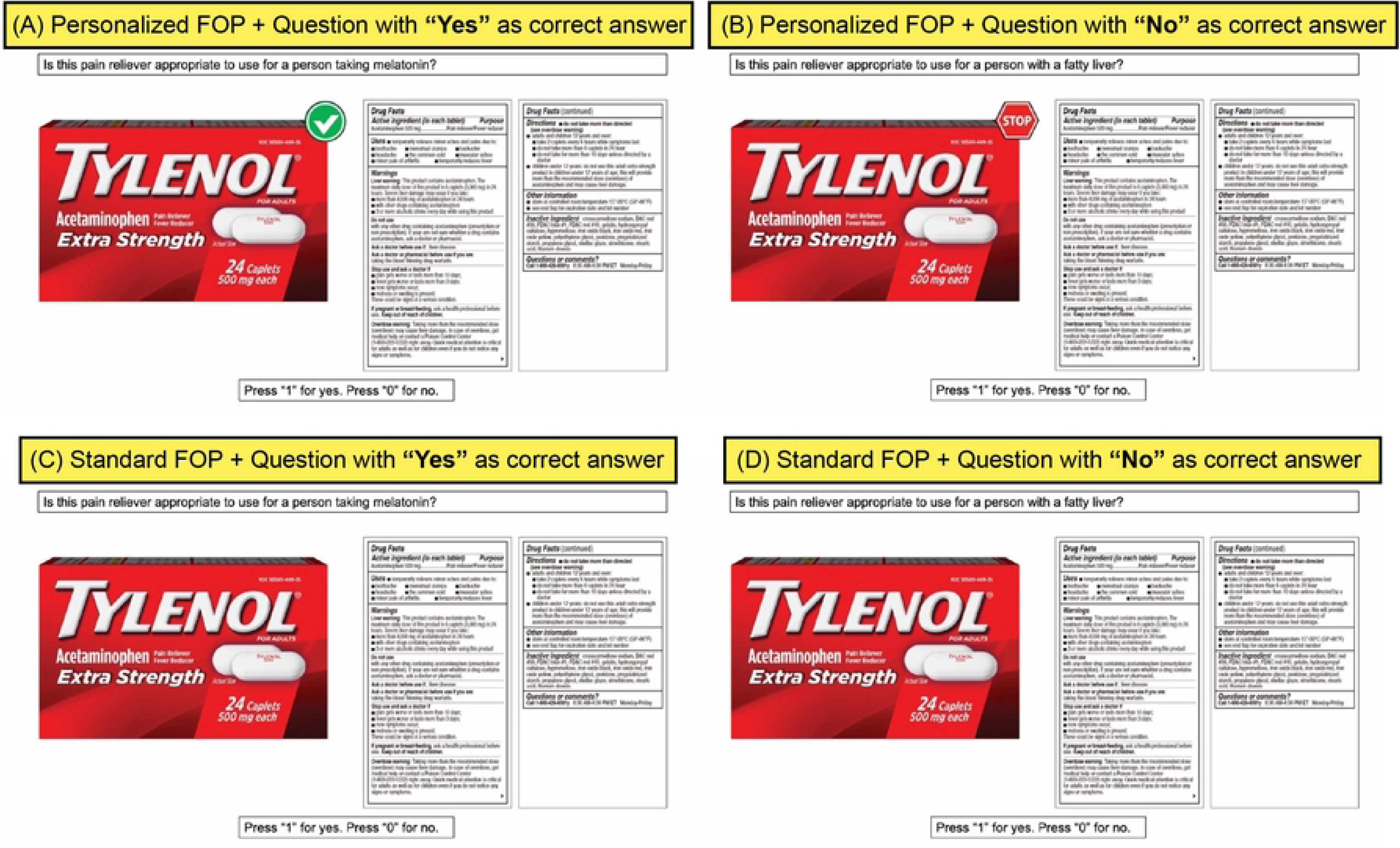
Single ingredient Acetaminophen containing product in four treatments. (A) Personalized labeling with “Yes” appropriate for use. (B)Personalized labeling with “No” appropriate for use. (C) Standard labeling with “Yes” appropriate for use yes. (D) Standard labeling with “No” appropriate for use.

One of the four possible treatments (answer type-appropriate for use/inappropriate for use x label design-personalized FOP/standard) of each brand/active ingredient (See Fig 2 for a single product) was randomly assigned to a single block of four possible experimental blocks (See Fig 3). The remaining three treatments were randomly assigned to each of the three remaining blocks. Trials that included questions about supplements (dummy trials) appeared between experimental blocks. This stratified-randomization pattern created a design of experiments where each brand/active ingredient only appeared once per block, and within each block the run order of the assigned treatments was randomized. This design meant that no active ingredient was presented in back-to-back trials, mitigating the likelihood of effects affiliated with short term memory. Neither practice nor dummy trials were included in the analysis of the data.

**Fig 3.**
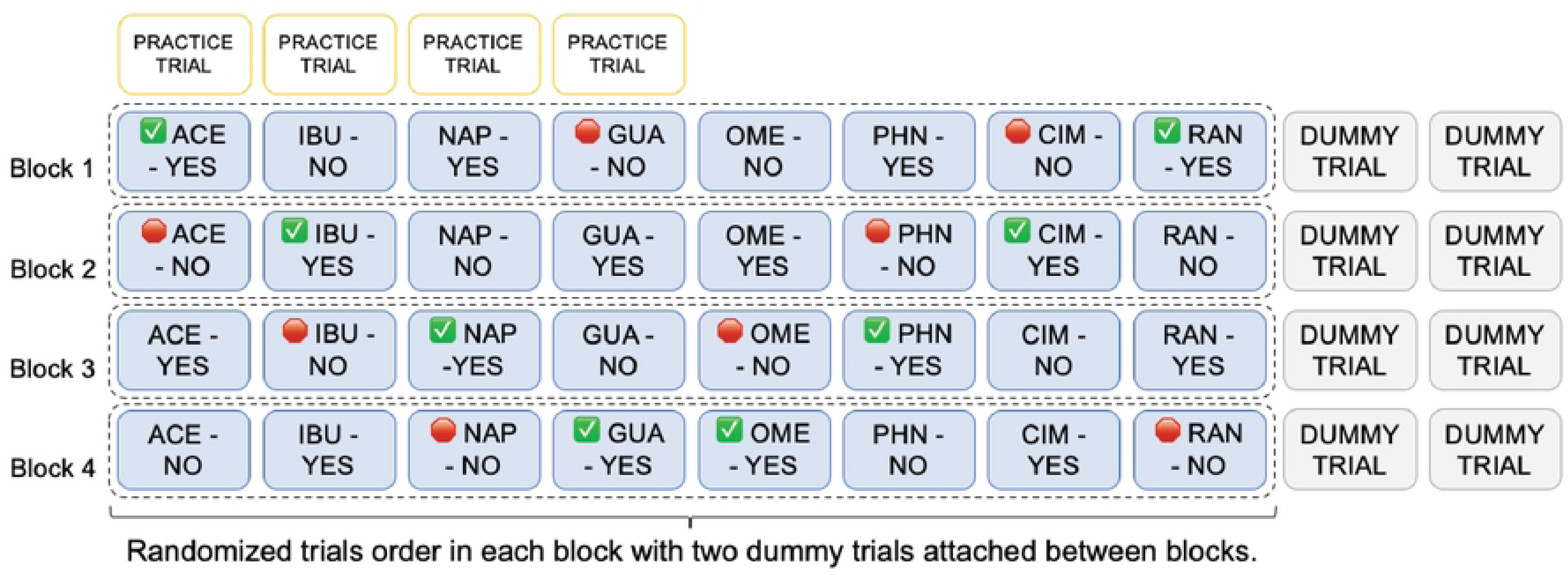
Stratified randomization scheme of the absolute judgement task.

Participants were instructed to assume that the person in the scenario had the condition/symptoms that the drug treats and answer whether or not the presented medication was appropriate for that person to take by depressing a “1” on the keypad for “yes/appropriate” and a z for no, not appropriate.

### 3. Statistical Analysis

A generalized, linear mixed model was fitted to assess the influence of the variables of interest on two outcomes (1) response accuracy, the probability of correctly answering the question for a given trial scenario, and (2) response time, the time to arrive at the correct response. For response accuracy, the binary data—correctly answered questions (yes/no) were interpreted in terms of probability (p) of correctly answering trial questions with logit transformation *ln*(*p*/1-*p*) to meet probability assumptions. Results of estimated means were back-transformed and displayed in terms of original target scale.

The response time data were checked for the validity of normality assumptions prior to statistical analyses. Residual plots and normal probability plots of the original data suggested an appropriate transformation, requiring a natural log transformation. Tukey’s method was used for minor non-constant variance and Satterthwaite’s method was used to adjust degrees of freedom. A generalized linear mixed model was then fitted to this natural log-transformed response time variable.

The predictor variables included in both of the final models were: group (concept educated/control), design layout (personalized label/standard), answer type (yes/no as a correct response), ingredients, education, ethnicity, sex, language, and age. All possible 2-way, 3-way interactions among group, design layout and answer type were also included in each model. Additionally, the following covariates were included in the final model: Participants were treated as random effects. All estimated means were back-transformed to the original scale of the dependent variable; in percentage for response accuracy, and in seconds for response time.

## Results

### 1. Participant characterization

Table 1 provides participant frequencies across demographic factors by group type for factors of interest for the entire study population.

### 2. Response accuracy

A total of 2,304 trials (72 participants × 32 trials) were analyzed in this absolute judgement test to examine data for effects on response accuracy. Participants provided answers correctly in 2,141 trials (92.9%) and incorrectly responded in 163 trials (7.1%).

A summation of the results from the statistical analysis is presented in Table 2. Significant main effects were identified related to design layout (p=0.002) and active ingredient (p=0.000) on response accuracy. Not surprisingly, a significant interaction was identified when design layout was crossed with group (p=0.003).

**Table 2.**
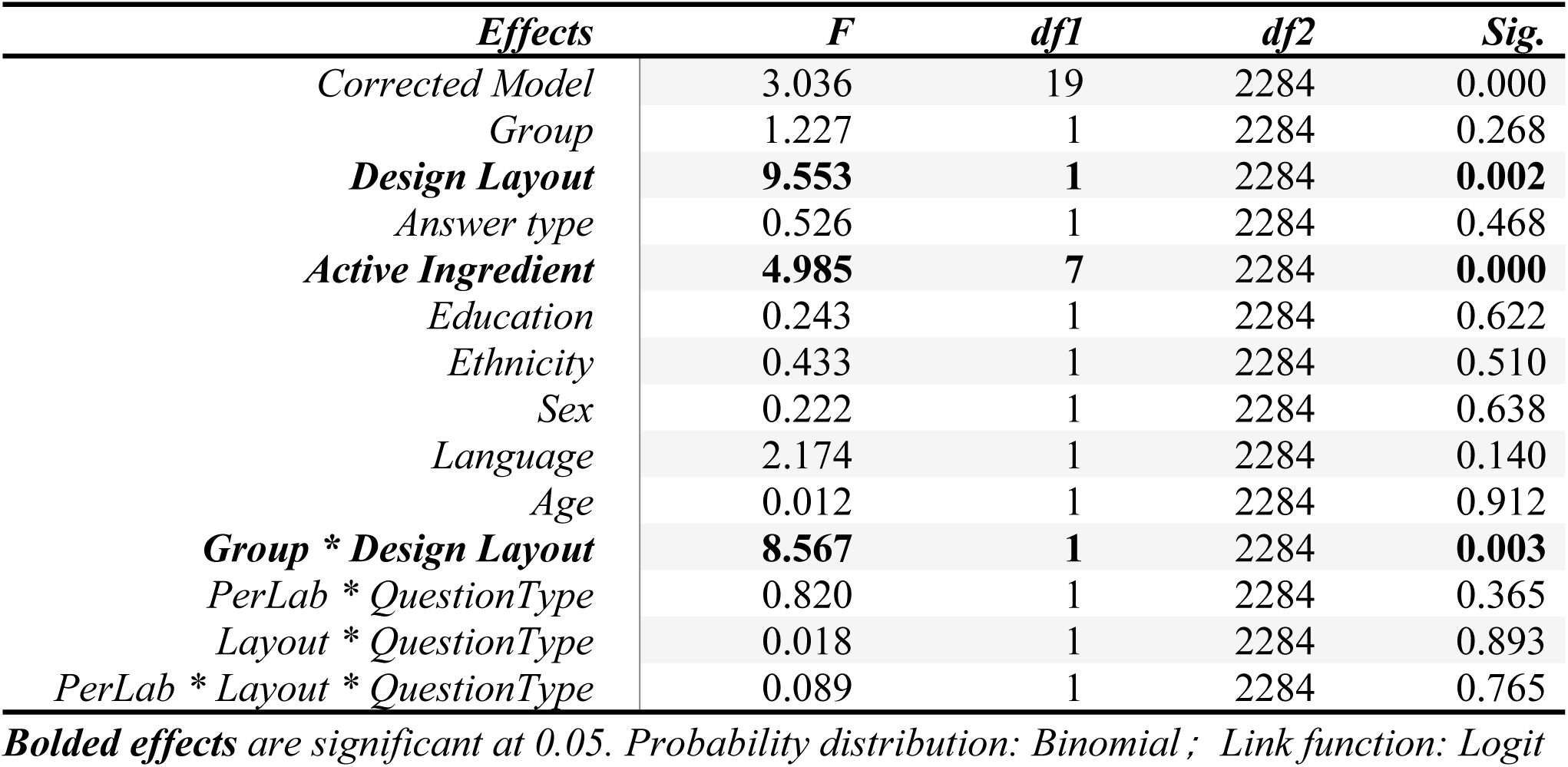
Tests of Fixed Effects on Response Accuracy.

Pairwise comparisons were conducted to interpret the significant 2-way interaction between group (concept-educated vs. control) and design layout (personalized labeling vs. standard) (See Fig 4). When the personalized labeling concept was introduced prior to the test, participants were significantly more likely to answer correctly in the trials with personalized labeling (ME=0.977, SE=0.007) as compared to the trials comprised of the standard formatting (ME=0.933, SE=0.017) (p=0.002). In contrast, no significant difference (p=0.898) in response accuracy was observed in the control group when trials utilizing personalized labeling (ME=0.946, SE=0.015) were compared with trials comprised of standard formats (ME=0.944, SE=0.015). Further, the accuracy in trials with personalized labeling responses shown to those in the concept educated group was significantly greater than any other combination of group and treatment at α =0.05 (See Fig 4).

**Fig 4.**
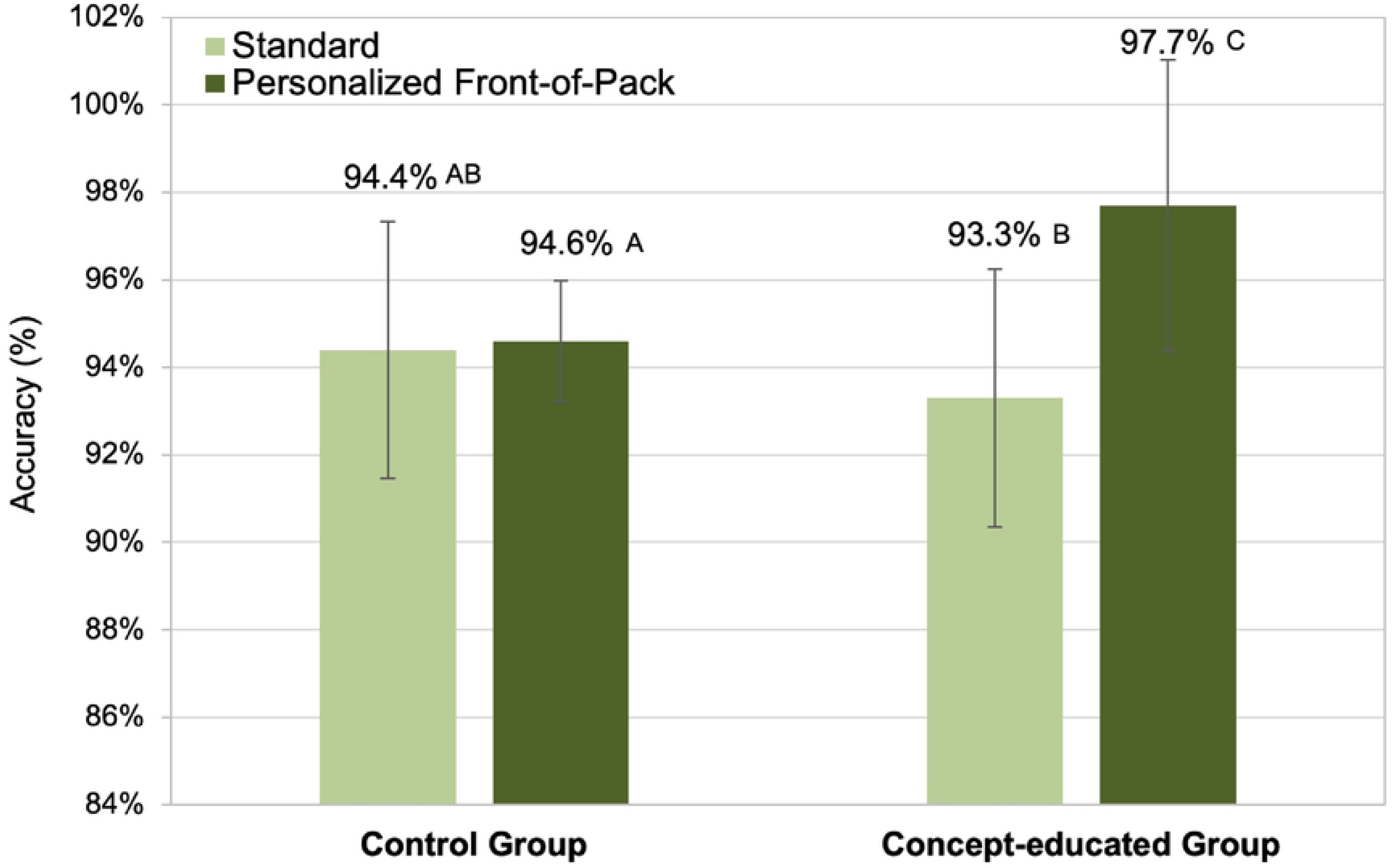
Estimated means of response accuracy (measure of effectiveness) on decision making by group and design layout. The error bars representing the standard error and the variables with differing letters indicate statistical significance at α=0.05

Within the covariates explored, the active ingredient was found to significantly affect the response accuracy (p<0.001). As shown in Table 3 and 4, Acetaminophen has the lowest estimated mean accuracy response. Comparisons indicated participants were significantly less accurate in responses for the trials that asked questions about acetaminophen (ME=0.909, SE=0.023), as compared to the trials involving responses associated with guaifenesin (ME=0.995, SE=0.023; p=0.001), or naproxen (ME=0.973, SE=0.010; p=0.018). Considering the popularity of acetaminophen usage in OTC medications and its risk, such as kidney failure from overdose (8), further study related to how active ingredient changes and patient characteristics such as previous use or drug familiarity influence information processing is recommended to gain further insight.

**Table 3.** Results of Estimated Means Accuracy Response Across Active Ingredients Tested.

| *Ingredient* | *Mean* | *Std. Error* | *95% Confidence Interval* |  |
| --- | --- | --- | --- | --- |
|  |  |  | *Lower* | *Upper* |
| *Ranitidine (acid reducer)* | 0.909 | 0.023 | 0.854 | 0.944 |
| *Phenylephrine (nasal decongestant)* | 0.958 | 0.013 | 0.922 | 0.977 |
| *Omeprazole (acid reducer)* | 0.948 | 0.015 | 0.909 | 0.971 |
| *Naproxen (pain reliever)* | **0.973** | **0.01** | **0.944** | **0.987** |
| *Ibuprofen (pain reliever)* | 0.916 | 0.021 | 0.863 | 0.949 |
| *Guaifenesin (cough suppressant)* | **0.995** | **0.004** | **0.978** | **0.999** |
| *Cimetidine (acid reducer)* | 0.909 | 0.023 | 0.854 | 0.944 |
| *Acetaminophen (pain reliever)* | 0.909 | 0.023 | 0.854 | 0.944 |

Continuous predictors are fixed at the following values: Age=34.42

**Table 4.**
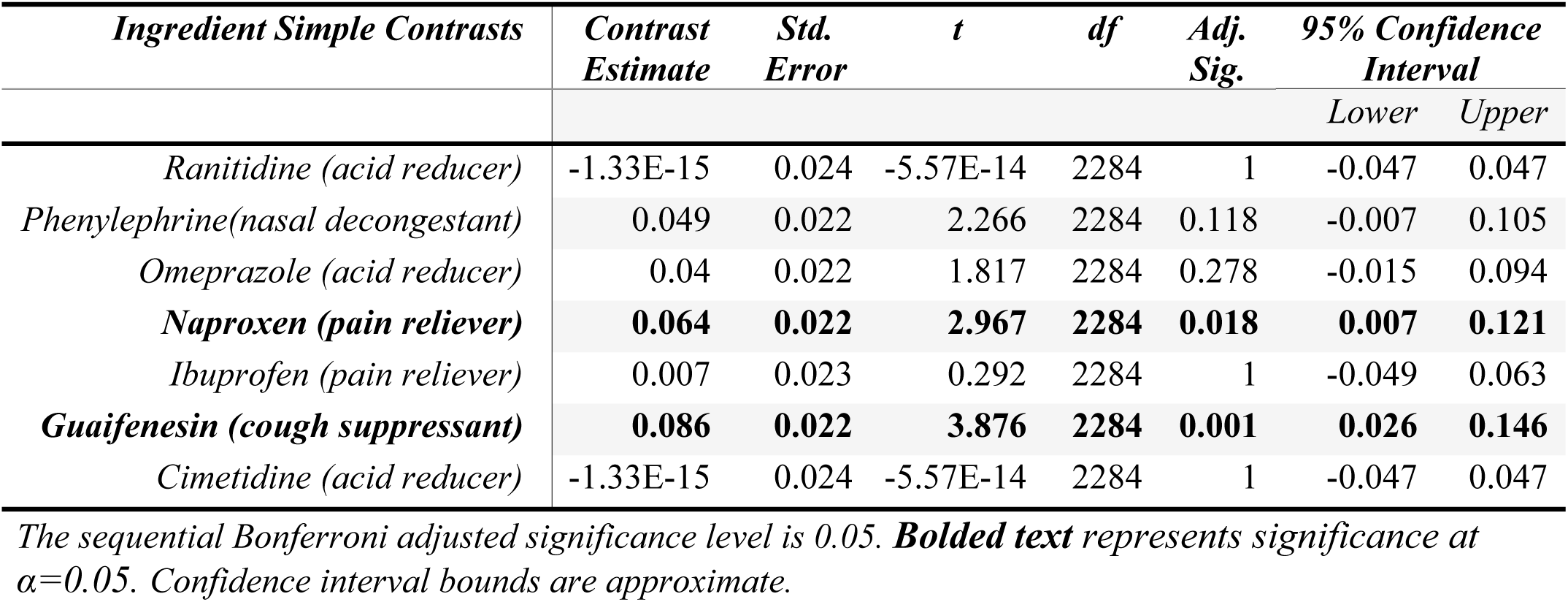
Simple Contrasts of Probability of Accuracy Comparing Each Active Ingredient to Response base (Acetaminophen, pain reliever)

### 3. Response time

While accuracy represents a measure of the design’s effectiveness, the time to arrive at a correct response can be considered a measure of the design’s efficiency (resources utilized to accomplish the stated goal, in this case, the correct response related to a drug’s appropriateness for use) (14). The response time for the 2,141 correctly answered test trials was used to analyze response time.

Results are presented in Table 5. Evidence was found to suggest significance for the main, fixed effects of group (p<0.001); design layout (personalized labeling vs standard labeling) (p<0.001); answer type (appropriate for use under a given scenario-yes/no) (p<0.001) and ingredients (p<0.001). Additionally, *all 2-way interactions* were found to statistically significantly impact time to correct response: specifically, group × design layout (p<0.001), group × answer type (p=0.000) and design layout × answer type (p<0.001). The 3-way interaction was also found statistically significant; group × design layout × answer type (p=0.015).

**Table 5.** Tests of Model Effects on Response Time for Correct Responses.

| *Source* | *F* | *df1* | *df2* | *Sig.* |
| --- | --- | --- | --- | --- |
| *Corrected Model* | 29.035 | 19 | 204 | 0.000 |
| **Group** | **21.629** | **1** | **65** | **0.000** |
| **Design Layout** | **111.276** | **1** | **2056** | **0.000** |
| **Answer Type** | **172.187** | **1** | **2057** | **0.000** |
| **Ingredient** | **15.819** | **7** | **2057** | **0.000** |
| *Education* | 3.8 | 1 | 65 | 0.056 |
| *Ethnicity* | 0.811 | 1 | 65 | 0.371 |
| *Language* | 2.711 | 1 | 65 | 0.105 |
| *Sex* | 0.13 | 1 | 65 | 0.719 |
| *Age* | 0.263 | 1 | 65 | 0.610 |
| **Group \* Answer Type** | **6.769** | **1** | **2057** | **0.009** |
| **Group \* DesignLayout** | **107.43** | **1** | **2056** | **0.000** |
| **Answer \*Design Layout** | **12.398** | **1** | **2057** | **0.000** |
| **Group \* Answer type \* DesignLayout** | **5.928** | **1** | **2057** | **0.015** |

**Bolded text represents significance at $\alpha = 0.05$**

To interpret the 3-way interaction that resulted in a significant effect on time to correct response (group (concept educated vs control) × design layout (personalized vs standard) × answer type (affirmative or negative response to appropriateness for a given scenario)), pairwise comparisons were analyzed. Fig 5 provides a visual assessment of this interaction.

**Fig 5.**
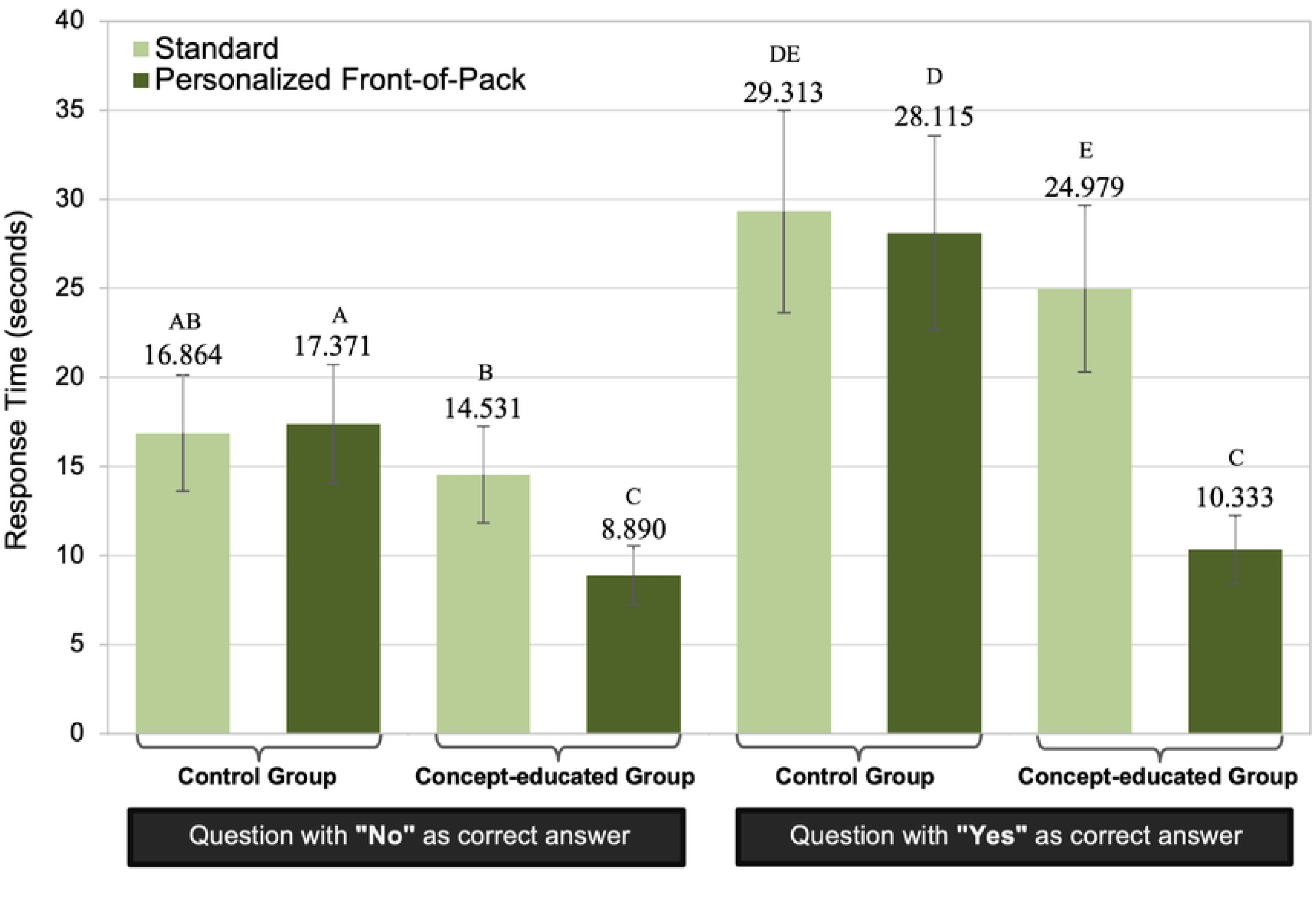
Estimated means of response time on correct decision making across group, answer type, design layout. The error bars representing the standard error and the variables with differing letters indicate statistical significance at α=0.05

For all comparisons related to answer type (appropriate yes and no) across the other variables- the concept educated group × personalized labeling; the concept-educated group × standard; control group × personalized; control group × standard), participants took statistically significantly longer to correctly respond to the affirmative questions-“Yes, appropriate” compared to the negative “not appropriate” as the correct response.

Data also supported the idea that people in the concept-educated group took significantly less time to make decisions than people in the control group, regardless of whether the correct response was affirmative, “Yes” (appropriate for the scenario) or negative, “No” (not appropriate for the given scenario). When collapsing the data across the label type (standard and personalized FOP), participants in the concept-educated group responded faster (p<0.001; ME=17.656, SE=1.825) than those in the control group (ME=28.714, SE=3.002) when the correct response was affirmative. Similarly, for trials with negative responses as correct, the concept-educated group responded faster (p<0.01; ME=11.711, SE=1.260) compared to the control group (ME=17.118, SE=1.791). These findings reflect that the concept-educated had shorter response times, regardless of the correct response type or the label type. Details relating to the pairwise comparisons for this analysis are shown in Fig 5.

The benefit of the personalized labeling strategy showed the most pronounced improvement when answering “Yes” after having been educated on the concept (ME=10.333, SE=0.979). The participants in this group made decisions significantly faster, an average difference of 17.782 seconds, than those in the control group (ME=28.115, SE=2.778).

To further explore the 3-way interaction, we also compared the effect of personalized labeling with the effect of standard labeling layouts within combinations of answer type and group. Results did not indicate evidence of significant difference (p=0.530) in response time to correct response when trials with personalized labeling trials were compared with trials employing the standard label were compared within the control group, the group not made aware of the concept (ME=28.115, SE=2.778 vs ME=29.313, SE=2.895 respectively). This was not the case for the concept educated group, where a significant difference for this comparison was noted. For the concept-educated group, more specifically, when trial questions were “yes” as correct answer, people spent 14.646 seconds less on average (p=6.66E-15) when the trials with personalized labels (ME=10.333, SE=0.979) were compared to responses resulting from the standard layout (ME=24.979, SE=2.388); similarly, when trial questions were “no” as the correct answer, people made decisions 5.641 seconds faster on average (p=1.53E-09) for trials which included a personalized label (ME=8.890, SE=0.842) compared with trials that included the standard layout (ME=14.531, SE=1.387).

Results related to correct answer response map onto target present (no, not appropriate for use—condition or behavior is contraindicated with a warning that is present in the DFL) and target absent (yes, appropriate for – no warning or behavior is contraindicated in the DFL) common in the literature. Specifically, these findings are consistent with published, visual search literature; it is well documented that reaction times are usually faster for target present than target absent searches (15).

## Discussion

We are in the era of the metaverse, a time where new technologies allow and enable us to connect and experience things in ways never imagined, impacting how we solve problems and make decisions. At the same time, dramatic changes are occurring in our approach to healthcare. The age of “personalized medicine” leverages the paradigm that the individualized and nuanced characteristics of a patient require interventions that are tailored to a patient’s specific molecular and physiological characteristics in order to yield optimized outcomes (16).

Within this context, we consider the state of OTC labeling. Extraneous information presented in the “one size fits all approach” to the DFL potentially interferes with a given patient’s ability to find information critical for *their unique condition and circumstance*. Catlin and Brass’(2018) review of OTCs and their observation that “the DFL by nature is static, non-customizable, text-based communication tool,“ that doesn’t “uniformly meet the needs of specific populations” (e.g. limited literacy, limited vision, older consumers or those with language barriers or preconceived notions related to the drug), suggests the need for a new archetype for labeling (3). Trends in emerging technology and increasing customization provide tremendous opportunity to rethink labeling practice to address identified shortcomings and challenges of the current DFL.

Our research goal was to provide objective evidence relating to the benefits of a theorized, personalized labeling strategy. Results suggest that this approach would enhance the effectiveness of decision making (accuracy – See Fig 4) as well as the efficiency (time to accurate decision – See Fig 5) over the current labeling standard (DFL), *when participants are aware of the new paradigm*. That said, the very nature of the application (a theorized app) would mean that patients that utilized it would be aware of it, and, as such, would not need to be educated related to the new paradigm.

The significant three-way interaction of group × label design × answer type catalyzed the need to explore the mediating effect of answer type (whether an affirmative or negative response as the correct response) on the response time of correct decisions. Analysis indicated that people took less time to find answers when “no” was the correct response as compared to those trials where “yes” as correct (See Fig 5). This finding is intuitive, as the questions were designed such that those with “no/not appropriate” as correct answers directly corresponded to key words from DFLs; in other words, there are visual targets present for the participant to find in the warning. This corroborates the work of others, who indicate that searches take longer when search targets are absent as compared to when targets are present in a visual search field (15,17–19). Using Tylenol, for example, we asked, “Is this pain reliever appropriate to use for a person with a fatty liver?”, where the keyword “fatty liver” corresponded to the “liver warning” and “ask a doctor before use if liver disease” on the DFL. Conversely, questions with an affirmative response/yes appropriate for use, would not contain directly correlated information in the DFL (visual target is absent. For the Tylenol example again, we asked “Is this pain reliever appropriate to use for a person taking melatonin?” In this case, the key words “taking melatonin” cannot be directly identified in the warning label. This finding is supported by the literature, which suggests that searching for an absent target (there is no warning present suggesting not to take this product) generally results in longer search times than target present searches (15,17–19). It should be noted, however, that the visual target is only absent in the case of the standard labeling as the personalized strategy provides a target in the form of a customized response tailored to the individual using the theoretical app.

It is somewhat surprising that people in the control group did not benefit from the personalized label – one might have thought that they would have realized that packages with a green checkmark were appropriate while those with a red stop sign were inappropriate. However, in our results they did not. In reality, people using the app that would be required to trigger the personalized response from the product would have to have some understanding of its purpose, so building awareness related to the same is not really an issue. From an experimental perspective, the fact that the personalized condition provided a benefited only in those educated about the personalization scheme provides confidence that it is the use of the scheme that led to benefits among the educated group

The final finding was a bit more unexpected. Namely, we note that the product itself (active ingredient) had significant impacts on both effectiveness (accuracy) and efficiency (time to correct decision). We hypothesize that this could be mediated by their familiarity with the drug, and note that in other studies conducted by our research team, people were actually more likely to make problematic decisions related to an OTC’s appropriateness for their own use when they indicated the active ingredient to be familiar to them (20).

## Data Availability

The data underlying the results presented in the study are available, in deidentified form upon request to the corresponding author.

## Acknowledgements

Thanks to the Communication Arts and Sciences SONA and the School of Packaging at Michigan State University for supporting this lab study. We also thank the Statistical Consulting Center at the College of Agriculture and Natural Resources, Michigan State University, for assisting with the statistical analysis. Thank you to the reviewers for their helpful feedback. This study was supported by the Healthcare Packaging Immersion Experience (HcPIE), a conference hosted by the research group. Additional support for publication fees and a small student stipend to assist with the publication process was provided by the Consumer Healthcare Products Association (CHPA). The content is solely the responsibility of the authors and does not necessarily represent the official views of HcPIE or CHPA.

